# Preschool-located influenza vaccination and influenza-like illness surveillance: an Italian pilot experience

**DOI:** 10.1101/2022.09.01.22279484

**Authors:** Antonella Amendola, Lucia Barcellini, Elisa Borghi, Silvia Bianchi, Nicolò Garancini, Chiara Nava, Alessandra Mari, Anna Sala, Clara Fappani, Maria Gori, Emerenziana Ottaviano, Daniela Colzani, Elia Mario Biganzoli, Elisabetta Tanzi, Gianvincenzo Zuccotti

**Affiliations:** Department of Health Sciences, Università degli Studi di Milano, 20142 Milan, Italy; EpiSoMI CRC-Coordinated Research Centre, Università degli Studi di Milano, 20133 Milan, Italy; Department of Pediatrics, V. Buzzi Children’s Hospital, Università degli Studi di Milano, 20154 Milan, Italy; Department of Clinical Sciences and Community Health, Università degli Studi di Milano, 20133 Milan, Italy; Department of Biomedical and Clinical Sciences, Università degli Studi di Milano, 20157 Milan, Italy

**Keywords:** children, school-located vaccination, live-attenuated influenza vaccination, self-sampling, saliva, health equity

## Abstract

Improving influenza vaccination coverage in young children is crucial in reducing the spreading of influenza viruses, thus indirectly protecting the general population. Mass campaign needs to engage all the communities within the target population, overcoming socio-economic and cultural barriers. School-located interventions in some countries were successful in increasing vaccine acceptability and overall immunisation rates.

Influenza-like illnesses (ILI) share several symptoms with the ongoing COVID-19 pandemic, posing an important challenge for both diagnostics and public health burden. Self-sampling using non-invasive saliva collection could represent a suitable tool for respiratory infection surveillance.

This pilot study was carried out in a very densely populated Italian area, Milan, characterised by a high percentage of non-Italian citizens, often hard-to-reach by preventive programmes. We started a vaccine campaign with live-attenuated influenza vaccine (LAIV), recently recommended in Italy for children aged 2-6, coupled with a saliva-based ILI surveillance system in preschools.

We enrolled 432 children from 5 schools in the Milan suburbs, 31.2% of which received the LAIV. Thanks to the undertaken multicultural approach, 59% of the vaccinated children were non-Italian citizens.

Despite the COVID-19 preventive measures, i.e., school closure upon positive cases, highly affected the programmed surveillance, all the saliva samples from ILI cases, self-collected by parents at home, resulted adequate for viral molecular testing.

With correction measures, this pilot experience could be broadened to a large population, simultaneously allowing for a widespread ILI surveillance and flu immunisation.

## 1 Introduction

The ongoing COVID-19 emergency has pointed out the need to reinforce measures to deal with a health crisis and to continue efforts in infectious disease prevention programs. Given COVID-19 overlapping symptoms with influenza-like illness (ILI) (1,2), influenza immunisation could ease the differential diagnosis and could be pivotal to implement public health strategies by policy makers (3). Influenza *per se* represents a major public health problem with important socioeconomic implications. The WHO recommends annual vaccination with inactivated or live attenuated vaccines, and outlines several risk groups for priority use including pregnant women, children, the elderly, persons with underlying medical conditions, and healthcare workers (4).

The 2020/21 flu season has been characterised by a dramatic reduction in the circulation of influenza viruses, likely due to the control measures put in place for COVID-19 and the limited importation of influenza cases into countries due to travel restrictions and border closures (5). Similarly, during the latest 2021/22 season, a lower number of influenza cases were reported compared to pre-COVID-19 periods, and fewer viruses were made available for characterization (5).

Nevertheless, with COVID-19 preventive measures and restrictions slackened, we can foresee an increase in influenza transmission and potential co-circulation of influenza viruses and SARS-CoV-2 with an additional burden on health services.

In light of these considerations, the WHO Strategic Advisory Group of Experts (SAGE) (6) recommends prioritising at-risk groups for influenza vaccination during the COVID-19 pandemic to ensure optimal influenza control and to reduce treatment in health care facilities that could lead to overload and increase the risk of exposure to SARS-CoV-2.

Despite current data indicating that children are not at increased risk of having severe COVID-19, they remain a priority group for influenza vaccination, particularly young children, because of their risk of severe influenza. Besides direct protection of the paediatric population, the primary spreaders of influenza, mass immunisation can provide indirect protection to the general population, reducing the public health impact. Yearly vaccine coverage is still below optimal rates for children in most countries (7,8). Parental hesitancy is one of the main reasons and results from several factors such as the concern about its effectiveness, the yearly booster, the wasted time visiting the provider office for a recommended but not mandatory vaccine, and, limited to pandemic years, the risk of SARS-CoV-2 exposure.

Several studies have demonstrated the feasibility and effectiveness of school-located influenza vaccination (SLIV) (9,10). Indeed, administering vaccines in school settings is a more efficient way to safely vaccinate large numbers of children. The United Kingdom implemented an immunisation program with live attenuated influenza vaccine (LAIV) nasal spray in primary care for children aged 2 to 3 years (11) and extended the school delivery model to additional year groups (12).

In Italy, until 2019, influenza vaccination was recommended for all individuals older than 65 years and for those with chronic diseases, regardless of age. Since the 2020/21 influenza season, vaccination was extended to all children aged 6 months to 6 years. To expand the vaccination coverage in the paediatric population, the Lombardy Region introduced for the first time in Italy the quadrivalent live attenuated influenza vaccine, licenced for 2-18 years old subjects, for children aged 2 to 6 years. The Department of Paediatrics of the University of Milan in conjunction with the local Health Authorities set up vaccination weekends in 6 different provider centres covering the metropolitan area of Milan. Nurses, healthcare assistants, medical specialists and resident doctors from the University of Milan were involved in the intervention. As a result, a total of 9292 children received intranasal vaccination, of whom 7675 were in the 2-6 years age group. Acceptability and safety data were also collected to assess possible barriers and tools that would help in planning the next vaccination campaigns. More than half of informed parents chose to vaccinate their children, and informal communication among parents in the same class group was the most effective tool in promoting the vaccination campaign. The LAIV proved to be non-invasive, simple, and convenient to administer. Most parents stated their intention to vaccinate their child again in the next flu seasons (13). As for many other health outcomes, disparities in influenza-related hospitalizations and deaths correlate with lower socioeconomic status (14,15). Therefore, strategies addressing these swathes of the population should be considered when designing targeted prevention programs such as vaccination campaigns. Moving in this direction, schools represent a unique setting and school-health programs have already proven to be one of the most cost-effective means available to improve both health outcomes and educational achievement (16,17). Moreover, for very young children and consequently for their parents, the school represents a familiar and trusted community environment, helping abolish barriers to vaccine access despite the socioeconomic status, and partially overcoming parental hesitancy due to the need of going to provider offices. In addition, influenza vaccination was demonstrated to improve school attendance and children wellbeing (18).

Children with ILI generally do not seek medical care, and not all ILI affected subjects are included in influenza surveillance systems (19). School-based surveillance can represent a valid option, but medical staff need to be present within the building to record symptoms and collect samples. Up to date, public schools in Italy do not fulfil this criterion. Oral fluids self-sampling or sampling under parents/legal guardians’ supervision might overcome this issue allowing virtually reaching all the pre-school and school population (20).

In the 2021/22 influenza season, a collaborative network between the University of Milan, Buzzi Children’s Hospital and the Milan Department of Education started a pilot SLIV experience preschools in Milan (Lombardy Region, Northern Italy), by immunising with nasal-spray LAIV and performing an ILI surveillance in the enrolled schools.

This article aims at describing the first school-located influenza vaccination campaign among pre-school children in Italy, and a pilot experience of ILI surveillance by saliva self-sampling, using schools as the connection point between children and parents, and investigators.

## 2 Context

In Italy, the central Authority defines the fundamental goals of the health system, and the Regions are responsible for organising and implementing healthcare services and preventive campaigns. In all the twenty Italian Regions, vaccinations are generally administered in dedicated provider centres, and school-located vaccination programmes - which have a long history in several countries - are not currently set up. Indeed, the last School-Located Vaccination (SLV) program set up in Italy in 1976 was the rubella vaccine campaign offered to prepubescent girls. Since the 1990s, with the introduction of the trivalent measles-mumps-rubella vaccine, the immunisation was extended to children of both sexes below the age of 2 years and administered in provider offices (21).

Influenza campaigns never took place in schools. For a successfully planned SLIV, the local context needs to be carefully evaluated.

Lombardy is the most populated region in Italy, counting one-sixth of the Italian population, and according to the Italian National Institute of Statistics (22), it ranked first for the total number of non-Italian citizens. Milan, its administrative centre, has a population of approximately 1.4 million inhabitants, including over 200,000 foreigners, mainly in the suburbs (23). Children aged 2-6 years old are about 55,000, of which 27% do not have the Italian citizenship.

Hence, any intervention in the school setting must encompass a communicative effort to reach all the school communities.

COVID-19 pandemic highly impacted on children’s school attendance, as institutes suffered several shutdowns, with opening/closure rules often revised according to viral circulation (24). In particular, during the study period and in pre-schools, up to January 2022, classes were forced to close for 10 /days after one positive case, then from February-April 2022, after four positive cases (25).

## 3 Community study design

### 3.1 Objectives

During the 2021/2022, we have set-up an innovative school-centred ILI surveillance and a preschool-located influenza vaccination program in the Milan area.

The purposes of this pilot study were: 1) to identify a suitable model of SLIV in local preschools to be implemented in the future to promote flu immunisation in children aged 2-6 years; 2) to set up a preschool-based screening to monitor acute respiratory infections, with a particular focus on influenza A/B and SARS-CoV-2.

### 3.2 Enrolment strategy

This was an observational surveillance study of ILI in preschool children who were offered influenza vaccination according to current recommendations and indications.

The study was proposed to 15 preschools in the Milan municipality by setting up informative meetings for local authorities, school principals, and teachers to promote the children’s enrolment. Informative materials, the informed consent form and the protocol received the approval by the local Ethical committee (protocol n. 0049030/2021).

LAIV (Fluenz Tetra, AstraZeneca) was offered to all healthy children, according to the current European Medicines Agency (EMA) authorization, and upon parent/legal guardian consent.

The lack of consent for LAIV was not an exclusion criterion for participating in ILI surveillance programme. Indeed, both vaccinated and unvaccinated children, whose parents/legal guardians signed for participation, could self-sample saliva for ILI differential diagnosis.

### 3.3 Sample collection and analysis

Lollisponge™ (Copan, Brescia, Italy) devices for saliva collection were delivered to parents together with detailed instructions on the collection procedure and on symptoms to be considered for ILI suspicion. In particular, ILI-related symptoms were sudden acute respiratory syndromes (cough, pharyngitis, nasal congestion) with onset of fever and/or headache, soreness, chills, sweating, and asthenia.

The LolliSponge™ is a non-invasive device that can be used to easily self-collect true saliva. The sampling is performed by keeping the sponge stick in the mouth for at least one minute without spitting or biting (26). Once duly soaked with saliva, LolliSponge™ can be kept at room temperature for up to 3 days until saliva processing.

In the study protocol, saliva was delivered by the parents to the child preschool and a pickup service was in charge of the shipment to the diagnostic laboratory the same day. SARS-CoV-2 test was performed within 24h, and the result referred according to the regional procedures. Briefly, true saliva was obtained by LolliSponge™ centrifugation for 1 minute at 450*g* (27), and stocked separately for SARS-CoV-2 and flu assays. SARS-CoV-2 detection was carried out by RT-PCR (Clonit COVID-19 HT Screen, CE-IVD for saliva). FluA and FluB presence was determined by a multiplex RT-PCR kit (Clonit Flu A + Flu B + RSV).

Any positive influenza sample was characterised by sequencing the genetic fragment coding for the HA1 subunit of the haemagglutinin (HA) protein, and by performing a phylogenetic analysis allowing to evaluate the similarity of the identified viral strain with the strains included in the 2021/22 vaccine.

In addition to saliva collection, parents were instructed to fill an e-form with any information about the respiratory illness, including the presence or not of concurrent systemic symptoms as well as information on occurrence of pneumonia, new onset or exacerbations of pre-existing cardio-respiratory conditions, hospitalizations, emergency room visits, and non-routine office visits and medication use within 30 days of illness start date. Data, both clinical and virological, were collected by investigators in a dedicated database. Paper questionnaire was also available for families not able to fill the online form.

## 4 Results

### 4.1 Vaccination campaign

Informative meetings for local authorities, school principals, and teachers were organised in September 2021 to promote the children’s enrolment. The recruitment procedure started at the beginning of October 2021, when pupils’ parents were informed about the study through thematic meetings, information leaflets, posters, and various school tools. Particular attention was paid to the engagement of non-Italian citizens, by involving cultural mediators and by setting up a multilanguage informative. Interested parents were asked to contact the study staff for any questions and to confirm child medical eligibility, and to sign the written informed consent.

A total of five public pre-schools were involved in the pilot project. The schools were based in the suburbs of Milan (Figure 1). The enrolled schools accounted for 432 children aged 2-6 years, 46% of which were not Italian citizens.

**Figure 1.**
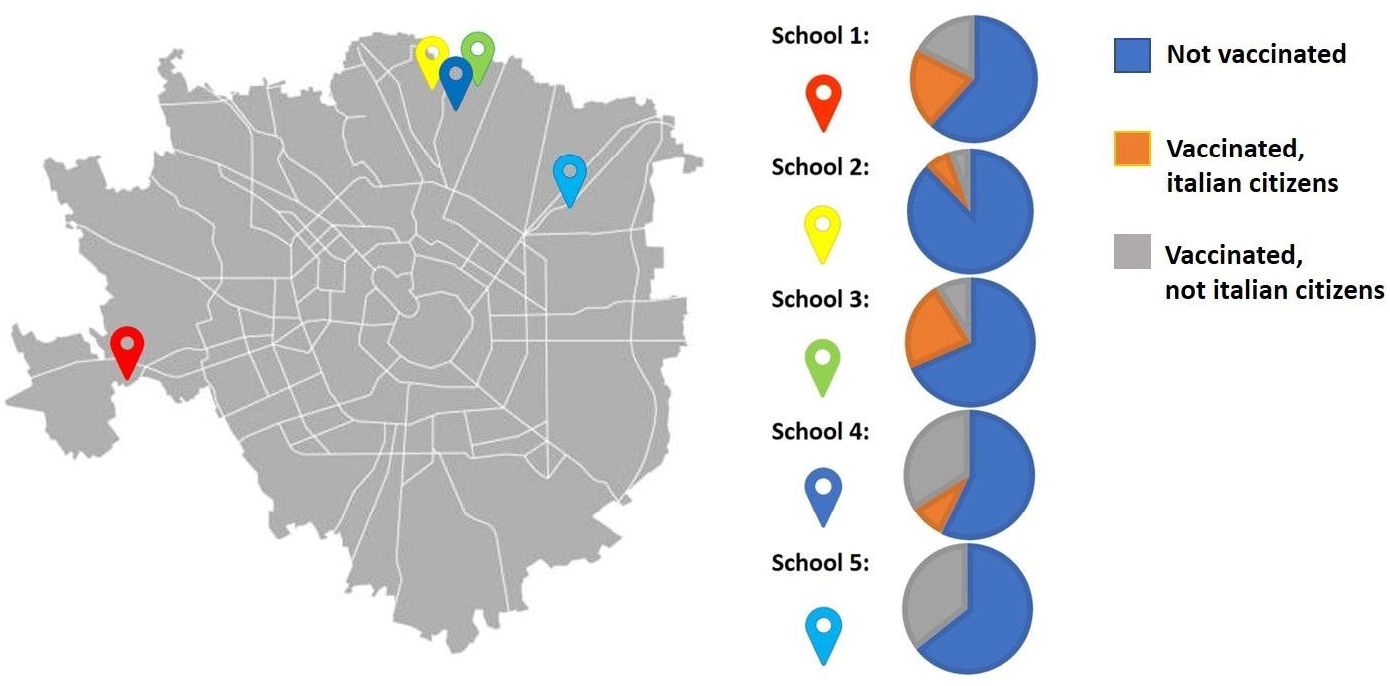
Milan urban area: school locations have been highlighted with coloured pins, and vaccination campaign results are reported as pie-chart for each school.

LAIV was offered to all healthy children without any contraindication, and immunisation was carried out in school settings by appropriately trained medical personnel who were part of the research team between November 9th and November 25th 2021.

The vaccination days were conducted directly within the schools, at the end of the classes or on the weekend, in order to encourage participation. With the collaboration of the teachers, the pupils were led one at a time with their parents into a dedicated room. Here, after obtaining informed consent, and once the suitability of the subject was checked, the vaccination was eventually carried out.

LAIV was administered as a divided dose sprayed into both nostrils. Immunised children were kept under observation for 20 minutes after vaccination to ensure their safety. Appropriate medical equipment and emergency medications, including epinephrine (1:1,000), were available on site in the event of an anaphylactic or other immediate allergic reactions.

A total of 135 (31.2%) pupils received the vaccination during the campaign, 59% of them were not Italian citizens (pie charts in Figure 1). No pupils experienced adverse reactions during the subsequent observation period.

### 4.2 ILI surveillance

Surveillance and follow-up took place from middle December 2021 (approximately 14 days after vaccination) until the end of April 2022.

In the study period, 19 saliva samples from 16 children were collected for ILI differential diagnosis. All the samples, i.e., saliva quality and quantity, were adequate for molecular testing, and 6/19 resulted positive for SARS-CoV-2. None of these presented with severe symptoms and need for hospitalisation. No differences were observed in the percentage of saliva samples from Italian and non-Italian children (6 and 10, respectively). In detail, 4 positive cases were detected in January 2022, one in February and one at the beginning of April 2022.

All the samples, despite SARS-CoV-2 positivity, were screened for Influenza A and B viruses. None of the samples collected in the surveillance tested positive for influenza viruses.

## 5 Discussion

To date, the vaccination programme described here is the first SLV reintroduced in Italy after almost 50 years. Notably, this is the first time that a SLV programme against influenza has been performed in Italy and a pioneer experience of SLV in pre-school children.

The overall participation in the immunisation campaign was good, considering possible absences due to COVID-19 cases in both pupils and parents, ranging from 11% to 42%, with more than half of the participants being of foreign origin. No adverse reactions were reported during the observation period, and thanks to the intranasal administration the procedure was well accepted both by children and parents.

The high participation of pupils of foreign origin in our campaign suggests that school-based campaigns can represent an effective strategy to reach families with language burdens. Indeed, informative meetings organised by the school with the presence of medical staff and cultural mediators were a crucial moment for rising parents’ engagement through a better understanding of the vaccination procedure and benefits.

Several improvements can be put in place to maximise participation. First, heterogeneity was observed in terms of participation reported among the five pre-schools included. The school with the lowest participation was the last enrolled in the project and the scarce adhesion may be partly explained by the short period between the information meeting and the scheduled vaccination session, which may not have allowed for adequate pass-on among parents who had not been able to attend the meetings.

Second, the campaigns were carried out on a single date for each school. The timing, at the end of classes or on the weekend, were decided based on school availability. No preliminary survey has been carried out among parents to check their preference and the presence of a parent/legal guardian was required in order to proceed with the vaccination. Preliminary surveys about parents’ preferences or offering more dates will contribute to the vaccination campaigns’ success.

Moreover, the pandemic period in which the campaign was carried out and the concomitant debate regarding the implementation of the SARS-CoV-2 vaccine among infants and children (28,29) may as well have affected the participation. In this regard, informative meetings offered a privileged time for a face-to-face discussion between parents and medical staff, and regardless of the final outcome in term vaccine acceptance, they represented a unique opportunity for health education.

Concerning the ILI surveillance, COVID-19 quarantine rules in preschools have highly hampered the program. Indeed, only nineteen saliva samples were brought to schools by parents for ILI differential diagnosis, because of the high class-closure rates. Despite the low number, the saliva-based testing was successful as all parents were able to collect an adequate amount of saliva, without professional assistance. This aspect is highly relevant, as self-sampling at home reduces children’s stress and anxiety and increases parents’ compliance. Saliva collection in young children is challenging as drooling, with or without straw/funnel is often unsuccessful (30) and they have not yet learned to spit. Oral swabbing could result in a scarce saliva amount not sufficient for multiple analyses, and cotton roll-based systems have choking hazard and is not recommended under 6 years of age (31).

The Lollisponge™ device is a lollipop-based sponge, provided with a detailed leaflet for collection procedures, that allows parents to successfully collect saliva at home. Saliva is self-preservative and now widely used for SARS-CoV-2 molecular testing. Butler-Laporte and co-workers carried out an exhaustive systematic review and meta-analysis on studies comparing SARS-CoV-2 nucleic acid amplification testing (NAAT) on both nasopharyngeal swab (NPS) and saliva; NAATs displayed similar specificity and sensitivity (32). Similarly, saliva showed similar performance for influenza viruses (33), resulting in a precious tool for respiratory infection surveillance in the paediatric population and to estimate the real-life effectiveness of LAIV.

Besides ILI surveillance, saliva self-sampling in school could guarantee a prompt SARS-CoV-2 diagnosis, allowing for a reduction of the turn-around-time and avoiding the exposure of medical staff to potential infectious risk. Moreover, our pilot experience revealed an even access to testing regardless of socioeconomic status, promoting equity in child health. An equitable access to preventive and diagnostic services is indeed a pivotal challenge of public health (34), especially in the management of epidemic and pandemic emergencies.

None of the collected samples was positive for influenza viruses. Of note, in the 2021/22 season, the influenza virus circulation has been mitigated by masks and social distancing, but a slight increase in activity was observed compared with the 2020/21 season. The epidemic peak was delayed to weeks 12 and 13/2022, in concomitance with COVID-19 preventive measures loosening (35,36). As we ended the ILI surveillance at the end of April 2022 (week 17), we may have missed some cases.

## 6 Conclusions

On the whole, the SLIV resulted in a good acceptability and feasibility, thanks to the intra-nasal administration, and was found to be well tolerated, partially helping in overcoming parents’ hesitancy. Our current experience, taking into account both positive results and limitations, could pave the way to a broader school-based vaccine campaign and ILI surveillance that eventually will support widespread access to health preventive measures, increasing vaccine coverage. At the same time, monitoring the viral circulation will enable us to estimate the impact of immunisation on reducing laboratory-confirmed influenza cases, complicated forms, and hospitalisation rates of children and their families.

## Data Availability

All data produced in the present work are contained in the manuscript

## 7 Conflict of Interest

The authors declare that the research was conducted in the absence of any commercial or financial relationships that could be construed as a potential conflict of interest.

## 8 Author Contributions

AA and GVZ conceptualized the project. AA and EBi made substantial contributions to the design of the study. LB handled the relationship between schools and University. LB, NG, CN, AM, and AS organized and carried out the vaccination campaign. CF, MG, EO, and DC carried out the ILI surveillance. AA, EBo, ET, and GVZ conceived the manuscript. EBo, AA, SB and LB wrote the first draft. All authors revised the manuscript and approved the final version.

## 9 Funding

This study was conducted with the unconditional support of AstraZeneca.

## 10 Acknowledgments

We thank children’s families for participating in this study, and school personnel that fully support the vaccine campaign. We thank the Lombardy Region for providing LAIV doses, and voluntary medical staff for vaccine administration.

## Notes

### Competing Interest Statement

The authors have declared no competing interest.

### Author Declarations

Ethics Committee of Milano Area 1 - ASST Fatebenefratelli-Sacco, protocol n. 0049030/2021, gave ethical approval for this work.

